# Worldwide Effectiveness of Various Non-Pharmaceutical Intervention Control Strategies on the Global COVID-19 Pandemic: A Linearised Control Model

**DOI:** 10.1101/2020.04.30.20085316

**Authors:** Joshua Choma, Fabio Correa, Salah-Eddine Dahbi, Barry Dwolatzky, Leslie Dwolatzky, Kentaro Hayasi, Benjamin Lieberman, Caroline Maslo, Bruce Mellado, Kgomotso Monnakgotla, Jacques Naudé, Xifeng Ruan, Finn Stevenson

## Abstract

**Background:** COVID-19 is a virus which has lead to a global pandemic. Worldwide, more than 130 countries have imposed severe restrictions, which form part of a set of non-pharmaceutical interventions (NPI)s. We aimed to quantify the country-specific effects of these NPIs and compare them using the Oxford COVID-19 Government Response Tracker (OxCGRT) stringency index, *p*, as a measure of NPI stringency.

**Methods:** We developed a dual latent/observable Susceptible Infected Recovered Deaths (SIRD) model and applied it on each of 22 countries and 25 states in the US using publicly available data. The observable model parameters were extracted using kernel functions. The regression of the transmission rate, *β*, as a function of *p* in each locale was modeled through the intervention leverage, *α_s_*, an initial transmission rate, *β*_0_ and a typical adjustment time, 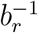.

**Results:** The world average for the intervention leverage, *α_s_* = 0.01 (95% CI 0.0102 - 0.0112) had an ensemble standard deviation of 0.0017 (95% C.I. 0.0014 - 0.0021), strongly indicating a universal behavior.

**Discussion:** Our study indicates that removing NPIs too swiftly will result in the resurgence of the spread within one to two months, in alignment with the current WHO recommendations. Moreover, we have quantified and are able to predict the effect of various combinations of NPIs. There is a minimum NPI level, below which leads to resurgence of the outbreak (in the absence of pharmaceutical and clinical advances). For the epidemic to remain sub-critical, the rate with which the intervention leverage *α_s_* increases should outpace that of the relaxation of NPIs.

## 1. Introduction

A novel corona virus, named COVID-19, was detected in the Hubei province of China in late December 2019 and rapidly spread resulting in a confirmed global pandemic by 11 March 2020^1^. As at 26 April 2020, the total number of infections has exceeded 3.1 million confirmed cases with more than 210 thousand deaths worldwide attributable to the effects of the virus^2^. A large, worldwide modeling effort is currently underway to improve health policy decision making with regards to the COVID-19 pandemic^3-7^. Many research groups and national response teams have looked into country specific intervention strategies^4,6-9^.

Recently, the total level of NPI stringency has been classified by the Oxford COVID-19 Government Response Tracker (OxCGRT) team and a nominal index measure has been defined for use by the wider international community. We address the use of this index measure to establish the degree and characteristics of control of the transmission rate of the virus within a representative sample of countries in the World and states in the United States of America.

## 2. Research in Context

### 2.1. Evidence before this study

Multiple teams have produced epidemic models which allow for the prediction of response to NPIs within their given locale. The typical complexity and bespoke nature of these models has meant that transferring insights from one country to another has been challenging. Comparing sufficient statistics (where they have been defined) amongst multiple other countries has not been done before.

### 2.2. Added value of this study

By comparing a wide variety of locales; we have shown a universality *of form* in response to changes in NPIs. Our modeling paradigm is succinct and requires only a few observable parameters to be measured in addition to what is presently done for successful control. These parameters allow for predicting not only *how much* effect a given NPI level has, but also *how long* we need to wait before we observe the effects.

### 2.3. Implications of the available evidence

The insight our research offers is a worldwide survey of the dynamics and characteristics various countries possess with regards to the control of the pandemic, through non-pharmaceutical interventions (NPI)s. Policy makers may now gradually control their local epidemic within their contextual situation without having to rely on the parameters derived from other country-specific intervention studies. This quantification allows policy makers to define safe limits of control.

## 3. Method

For a fair comparison of the effect of interventions, carefully curated, comprehensive and frequently updated data is a requirement. Data was, therefore, sourced from the Johns Hopkins COVID-19 data repository because it fulfilled both the due diligence and frequency of updating needed for this research.^10^

Using this existing data up to and including 26 April 2020, multiple SIRD with latent dynamics and error propagation models (see Appendix B for full details) were employed. The chosen modeling method is simpler than others used in this pandemic.^7^ This is a feature since it is ‘mind-sized’ and has a number of sufficient statistics needed to understand the response to control. An initial cohort of 23 countries and 25 states within the United States of America were chosen to be as representative and diverse as possible.

Levels of control were measured by the Oxford COVID-19 Government Response Tracker (OxCGRT) stringency index.^11^ This index, denoted *p* in our work, takes values from 0 to 100 and is indicative of the severity and number of discrete measures that have been taken to try to curb the spread of COVID-19.^11^

At the time of writing; America did not have a stringency index published. Incorporating their reported NPIs into the OxCGRT framework described fully in Appendix A.

Daily parameter estimates of the models were extracted from the data using a least-squares framework and candidate kernel functions were regressed on the daily parameter estimates. These kernel functions represent the time domain response of a model parameter to a given level of control and are based on a dynamic hypotheses that additive effects of random variables contribute to the observed effects of intervention. ^12^

This sort of exponential behaviour will cease to be representative if there is a systematic breakdown in roll-out of the interventions (so called ‘fat tails’).^13^ If such a systematic breakdown were to happen, our proposed kernel functions would cease to be valid.

### 3.1. Model

The characteristics of the present pandemic are atypical in that infected individuals may be asymptomatic and infectious. ^14^

To this end, we developed a dual observable/latent SIRD model to capture the essential features of the present COVID-19 outbreak and allow for clear interpretation with regards to control and decision making. This model is depicted in Figure 1 and has several key parameters which completely characterise the behaviour.

**Figure 1:**
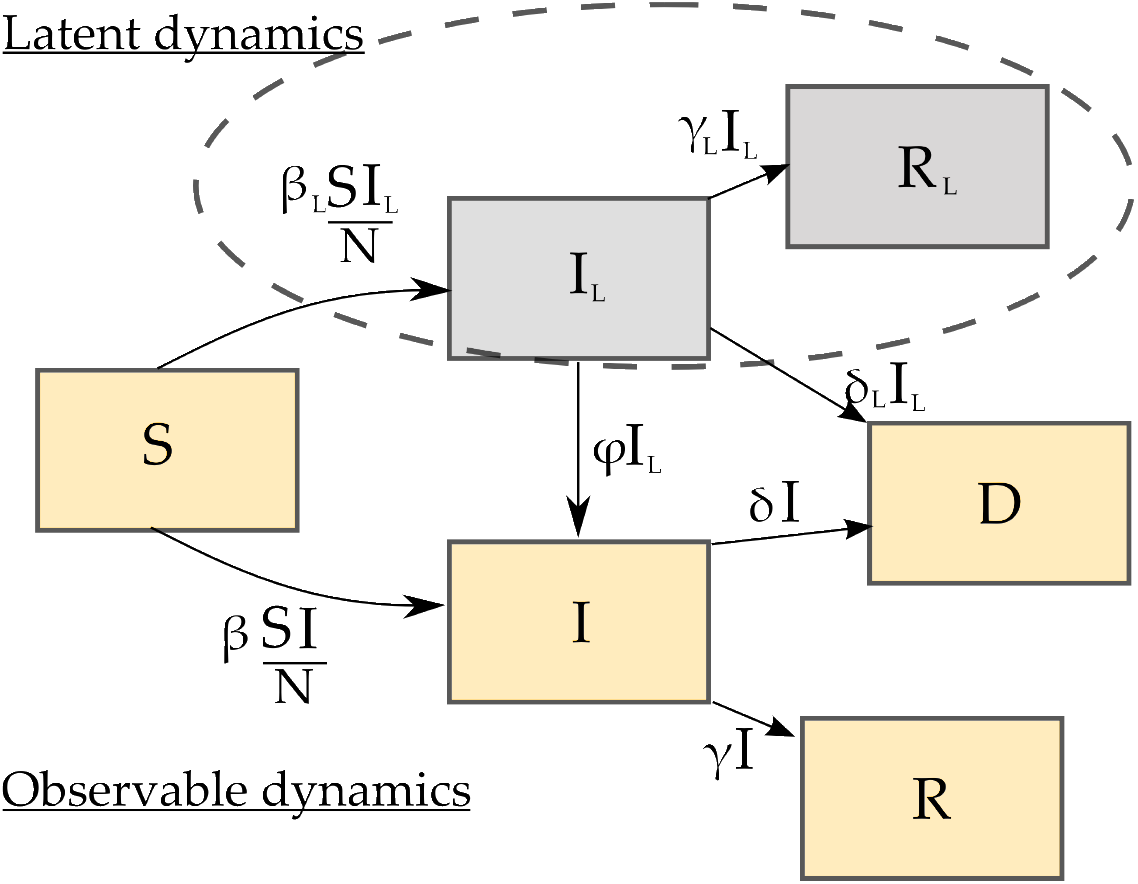
SIRD model with Unobserved Latent Dynamics.

The latent dynamics are essential to capture the following clinical properties of COVID-19:

1. Asymptomatic cases are infectious^15,16^.
2. It is possible for deaths to be overstated since positive tests are not a requirement in some locales for establishing the basic underlying cause of death in this pandemic; reasonable clinical judgement is^17^. In addition, overstatements in clinical cases may be due to the highly infectious nature of the virus; patients with severe comorbidities may not necessarily die with the virus as the basic underlying cause even though it is likely to be present in the patient if they are admitted to a facility with many active COVID-19 cases.
3. There are early reports that the latent prevalence (using a seroprevalence study) may be as high as 14% of one of the most densely populated cities in the World^18^.
4. There have been serious calls for protection of front-line health care workers as a result of early experiences with the epidemic^19,20^. The nosocomial infections within hospital care providers as a group may be non-negligible, though for the large scale model we are considering, this group has not been partitioned separately.

The observable (directly measurable) dynamics capture the standard features of susceptible portions of the population becoming infected and then either recovering with perpetual immunity or dying.

There is evidence that relapse is possible, where recovered patients would become susceptible again^21^. This model variation has not been explicitly considered because do not know the significance of this yet; it is possible that these are people who were not fully cured^21^, they may have been reinfected with a different strain^16^, the reverse transcription polymerase chain reaction (RT-PCR) test can give a positive result due to extracellular RNA, shedding of non-viable virions etc.

Of note is that the implicit assumption with this approach is that there is a single viral strain which is being modelled. If there are multiple strains, each of which is observable and differentiable through specialised testing then refinements on the labelling and parameters are possible.

### 3.2. Observable Dynamics

Under the stated clinical conditions the transmissions between known infected patients, *I*, and susceptible individuals (in the broadest sense possible) should be rare. The typical disease progression time frame^14,22^ implies that latent infections leading to deaths should be rare, it is presumed that patients with severe symptoms are brought in for treatment. Under these conditions the latent dynamics in the model simplify with *δ_L_* ≈ 0 and *β* ≈ 0 and this model simplification is used throughout.

What is observed then, in terms of increase in measured infections *I*, is achieved strictly through *ϕI_L_*. These are the detected infections and dependent on the detection rate, *ϕ*.

### 3.3. Daily observed infection rate, ϕ

An important component for estimating *ϕ* is the number of positive individuals who do not know their status and are asymptomatic. Random testing has proven useful in this regard in Iceland^16^ and New York^18^.

Reliable estimates of asymptomatic cases due to either extensive randomised testing or exhaustive testing in closed populations are presented in Table 1. Unfortunately, the number of asymptomatic cases are approximately 50% of all positive COVID-19 patients. The fraction of asymptomatic cases in enclosed populations, the number of patients with antibodies present (due to randomised population seroprevalence studies) and patients with symptoms who are sub-clinical and do not warrant admission and formal testing will also all influence the rate measured by *ϕ*.

**Table 1:** Results from various studies regarding the number of asymptomatic cases amongst those testing positive for COVID-19.

| Study | Asympt. positive tests |
| --- | --- |
| Diamond Princess<sup>14</sup>[n=4,062] | 55% (95% CI 52%-59%) |
| Vo Italy<sup>23</sup>[n=3,300] | 50% - 75% |
| Japanese nationals evacuated<sup>24</sup> [n=565] | 31% (95% CI: 7.7% - 54%) |
| Nursing home, Washington<sup>25</sup> [n=23] | 43% (95% CI 26% - 63%) |
| Iceland (Open Invite)<sup>16</sup> [n=10,797] | 41% (95% CI 32% - 52%) |
| Iceland (Random)<sup>16</sup> [n=2,283] | 54% (95% CI 30% - 77%) |
| Northern Italy blood donors<sup>26</sup> [n=60] | 66% (95% CI 54% - 77%) |

### 3.4. Kernel functions

The roll-out of any country-wide control measure is subject to random variations in timing and levels of civil compliance dependent on the execution plan and the populace’s disposition. This was modeled with the kernel function, conditioned on the control measure *p*:

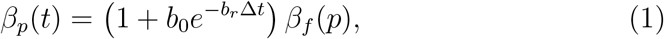

where *β_f_* (*p*) is the asymptotic value of the observed daily transmission rate, *b_r_* is the typical adjustment rate and *b*_0_ is used to model the initial transmission rate before the control *p* is applied. A characteristic plot is depicted in Figure 2.

**Figure 2:**
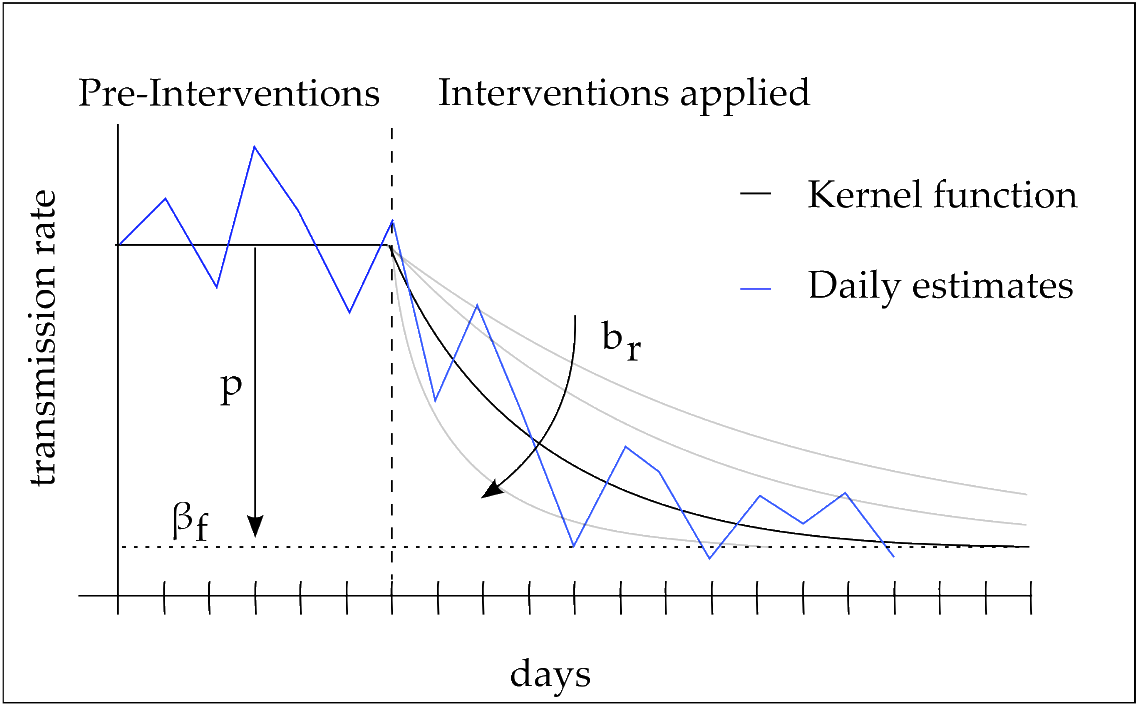
The transmission rate kernel function depicting the typical adjustment rate, *b_r_* and final transmission rate *β_f_*. The control index *p* affects the total reduction in transmission.

As a starting point, it is hypothesised that the typical adjustment rate *b_r_* is characteristic of a nation and that *β_f_* (*p*) is dependent on the stringency of control *p* through:

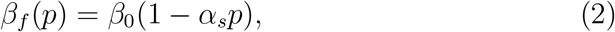

where *α_s_* is the intervention leverage. In the absence of data on the population dynamics during withdrawal of some NPI’s, our model is conservative and uses the same typical adjustment rate for the withdrawal of NPI’s as for the addition of NPIs.

The kernel function for *γ*(*t*) is:

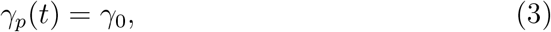

regardless of control. The clinical and physical justification for this model is that the inherent properties of the recovery process do not change with non-pharmaceutical control, assuming the hospital system has not been overwhelmed yet.

The kernel function for *δ*(*t*) is:

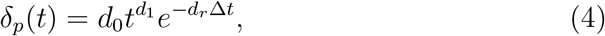

which is the result that would be theoretically expected from a cascade of first order processes^27^. The temporary increase in this parameter is due to the reduced number of severe infections which will artificially raise it since 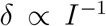, see (B.7). This function captures the transients inherent in the daily death count as a result of intervention only.

### 3.5. Control

The capacity of countries to properly address the risk that COVID-19 poses is varied; with less than half being fully positioned to prevent, detect, and respond appropriately^28^. The World Health Organisation identifies the need to reduce human-to-human transmission of the virus using numerous public health measures. ^2^

The effect of these various control policies on our latent model are depicted in Figure 3. This Figure clearly demonstrates the value of the proposed methodology; one may holistically view the epidemic and various policy actions in a single framework and be able to anticipate the consequences.

**Figure 3:**
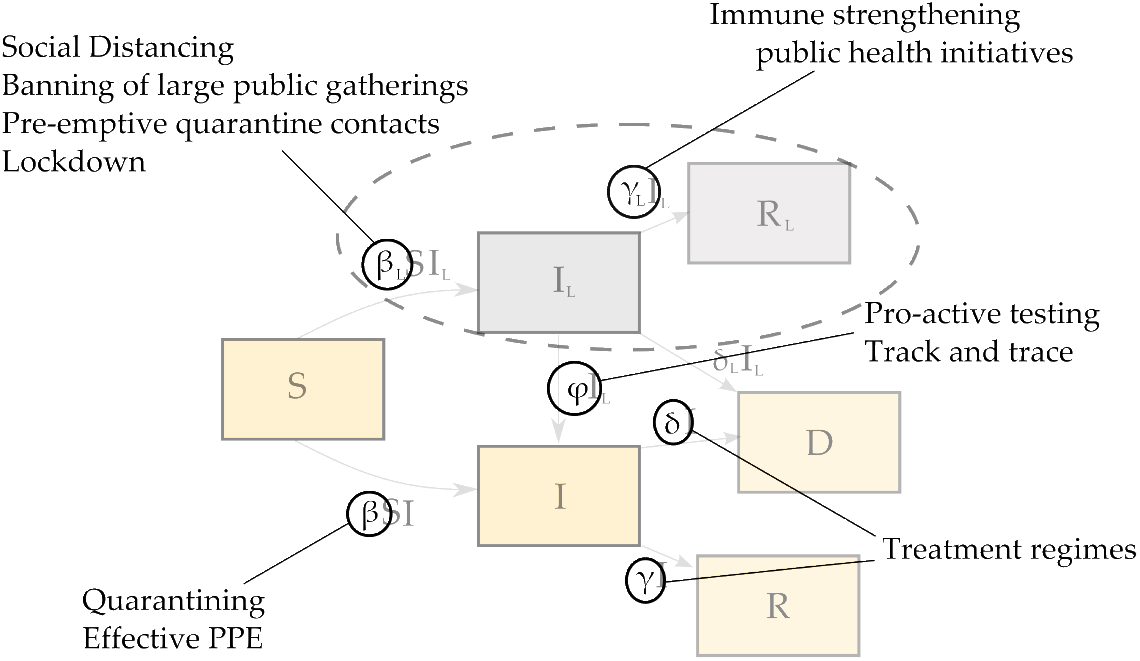
The Effects of Various Interventions on the Model

A method to measure the overall level of control is described next.

### 3.6. NPIs and the OxCGRT Stringency Index

The OxCGRT has developed a valuable database for comparing countries response strategies.^29^ The database contains the following levels of control (coded using ordinal numbers) and timing for 139 countries:

1. S1 - School closure
2. S2 - Workplace closure
3. S3 - Cancel public events
4. S4 - Close public transport
5. S5 - Public information campaign
6. S6 - Domestic travel bans
7. S7 - International travel bans

Also included in this data set is a Stringency Index, *p* in our notation, which provides a single number that captures the overall level of intervention implied by combinations of the ordinal numbers S1-S7. The Oxford stringency index is calculated using a weighted average of the above seven non-pharmaceutical interventions ^30^.

As an example of the use of this index, the overall intervention level is depicted in Figure 4, plotted against the number of days since the first reported COVID-19 case.

**Figure 4:**
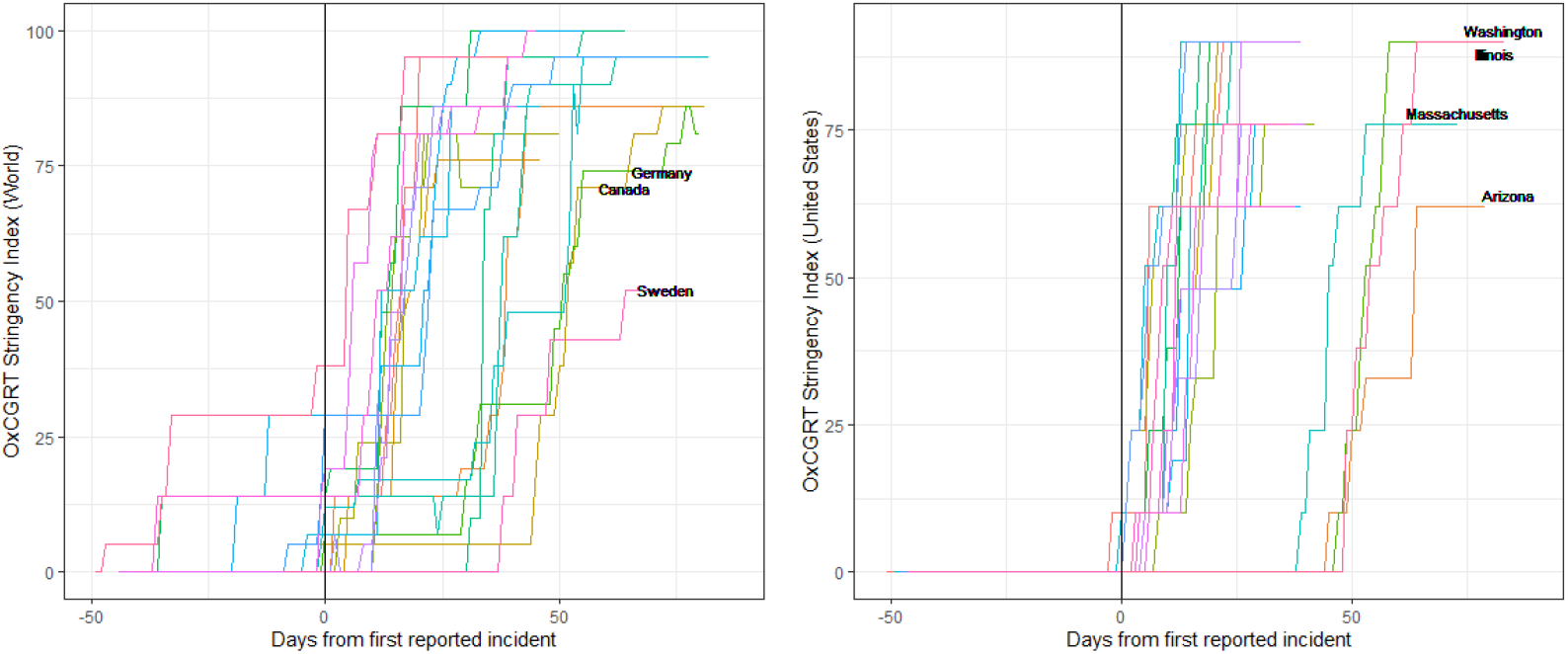
Timing and Severity of NPIs in our set of representative countries and US

## 4. Results

The results, using the foregoing methodology are shown in Table 2 for the selected countries in this study and in Table 3 for selected States of the US. Results are expressed in terms of *γ*_0_, *d*_0_, *β*_0_, 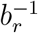, *p_max_* and *α_s_* in Table 2. Table 3 does not display *γ*_0_, as the data for the number of recoveries has not been reported in the US since late March^10^. The first salient feature of Tables 2 and 3 is that the evolution of the pandemic in different countries share strong similarities. Some variations are observed in the parameters *γ*_0_, *d*_0_, *β*_0_, where 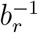 is subjected to features of the data most likely to specifics in the reporting. Of note though is the consistency of *α_s_*.

**Table 2:** Results for various representative countries around the World. Results are given in terms of *γ*_0_, d_0_, *β*_0_, 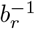*, p_max_* and *α_s_* (see text). Parameters have been obtained with data up until April 26 2020.

| Country | $\gamma_0$ | $d_0$ | $\beta_0$ | $b_r^{-1}$ [days] | $p_{max}$ | $\alpha_s$ |
| --- | --- | --- | --- | --- | --- | --- |
| Austria | 0.045 | 0.00342 | 0.26 | 7.15 | 95 | 0.010 |
| Belgium | 0.019 | 0.00070 | 0.24 | 19.12 | 86 | 0.012 |
| Brazil | 0.034 | 0.01670 | 0.39 | 2.97 | 76 | 0.009 |
| Canada | 0.026 | 0.00333 | 0.31 | 13.53 | 86 | 0.010 |
| Chile | 0.036 | 0.00150 | 0.30 | 5.74 | 81 | 0.009 |
| Ecuador | 0.006 | 0.00004 | 0.42 | 2.89 | 100 | 0.009 |
| Egypt | 0.027 | 0.00058 | 0.15 | 9.18 | 100 | 0.005 |
| France | 0.023 | 0.00012 | 0.41 | 8.32 | 95 | 0.009 |
| Germany | 0.042 | 0.00258 | 0.28 | 10.50 | 86 | 0.011 |
| Ireland | 0.015 | 0.00372 | 0.34 | 23.46 | 86 | 0.012 |
| Israel | 0.018 | 0.00003 | 0.27 | 8.89 | 100 | 0.009 |
| Italy | 0.020 | 0.00107 | 0.24 | 10.26 | 95 | 0.010 |
| Morocco | 0.011 | 0.0021 | 0.25 | 16.58 | 86 | 0.010 |
| Netherlands | 0.001 | 0.00050 | 0.22 | 16.21 | 86 | 0.012 |
| Peru | 0.051 | 0.00163 | 0.33 | 1.49 | 86 | 0.007 |
| Portugal | 0.002 | 0.00045 | 0.31 | 8.28 | 100 | 0.009 |
| South Africa | 0.018 | 0.00050 | 0.28 | 2.19 | 100 | 0.008 |
| South Korea | 0.032 | 0.00031 | 0.07 | 16.24 | 81 | 0.012 |
| Spain | 0.037 | 0.00006 | 0.34 | 11.96 | 95 | 0.010 |
| Sweden | 0.003 | 0.00002 | 0.12 | 38.09 | 52 | 0.019 |
| Switzerland | 0.036 | 0.00045 | 0.46 | 8.12 | 81 | 0.012 |
| Turkey | 0.012 | 0.00047 | 0.64 | 14.41 | 95 | 0.011 |
| UK | 0.001 | 0.00027 | 0.23 | 14.37 | 71 | 0.013 |

**Table 3:** Results for various representative states within the United States of America. Results are given in terms of *γ*_0_, *d*_0_, *β_0_*, 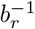, *p_max_* and *α_s_* (see text). Parameters have been obtained with data up until April 26 2020.

| Country | $d_0$ | $\beta_0$ | $b_r^{-1}$ [days] | $p_{max}$ | $\alpha_s$ |
| --- | --- | --- | --- | --- | --- |
| Alabama | 0.00074 | 0.40 | 9.01 | 90 | 0.011 |
| Arizona | 0.00009 | 0.34 | 5.88 | 62 | 0.014 |
| California | 0.00093 | 0.23 | 10.56 | 90 | 0.010 |
| Colorado | 0.00011 | 0.30 | 1.41 | 90 | 0.011 |
| Connecticut | 0.00057 | 0.54 | 12.19 | 76 | 0.013 |
| Georgia | 0.00015 | 0.35 | 15.53 | 76 | 0.013 |
| Illinois | 0.00036 | 0.39 | 9.20 | 90 | 0.010 |
| Indiana | 0.00127 | 0.33 | 5.99 | 90 | 0.010 |
| Louisiana | 0.00044 | 0.63 | 5.59 | 90 | 0.011 |
| Maryland | 0.00282 | 0.34 | 10.66 | 90 | 0.010 |
| Massachusetts | 0.00251 | 0.25 | 8.98 | 76 | 0.011 |
| Michigan | 0.00102 | 0.97 | 8.11 | 90 | 0.011 |
| Missouri | 0.00003 | 0.58 | 3.45 | 76 | 0.012 |
| Nebraska | 0.00079 | 0.23 | 3.12 | 62 | 0.013 |
| New Jersey | 0.00002 | 0.50 | 8.20 | 90 | 0.011 |
| New York | 0.00006 | 0.45 | 10.71 | 90 | 0.011 |
| North Carolina | 0.00131 | 0.39 | 9.49 | 90 | 0.010 |
| North Dakota | 0.00022 | 0.44 | 10.04 | 48 | 0.018 |
| Ohio | 0.00066 | 0.48 | 15.91 | 90 | 0.011 |
| Oklahoma | 0.00025 | 0.44 | 4.75 | 76 | 0.012 |
| Pennsylvania | 0.00005 | 0.38 | 6.81 | 90 | 0.010 |
| Texas | 0.00009 | 0.35 | 8.17 | 76 | 0.012 |
| Utah | 0.00049 | 0.46 | 4.86 | 62 | 0.015 |
| Virginia | 0.00135 | 0.34 | 5.38 | 76 | 0.011 |
| Washington | 0.00035 | 0.19 | 5.50 | 90 | 0.010 |

### 4.1. Ensemble variation of intervention leverage, α_s_

With a formal treatment, the following ensemble characteristics are observed with regards to the intervention leverage across all locales: the ensemble average intervention leverage is given by *α_s_* = 0.01 (95% CI 0.0102-0.0112) and the ensemble standard deviation is given by 0.0017 (95% C.I. 0.0014 - 0.0021). These results depend on the respective marginalisation over a Gaussian noise process, a flat prior probability for the mean and a Jeffrey’s prior for the variance.^12^ Note that there were two outliers that were removed from this calculation, Sweden and North Dakota and they are left for discussion.

Using the values from Tables 2 and 3, the expected control surfaces which map stringency level *p* to transmission rate *β* are depicted in Figures 6 and 7 respectively. There are clearly a few outliers which are left for discussion, but the overall behaviour is remarkable.

**Figure 5:**
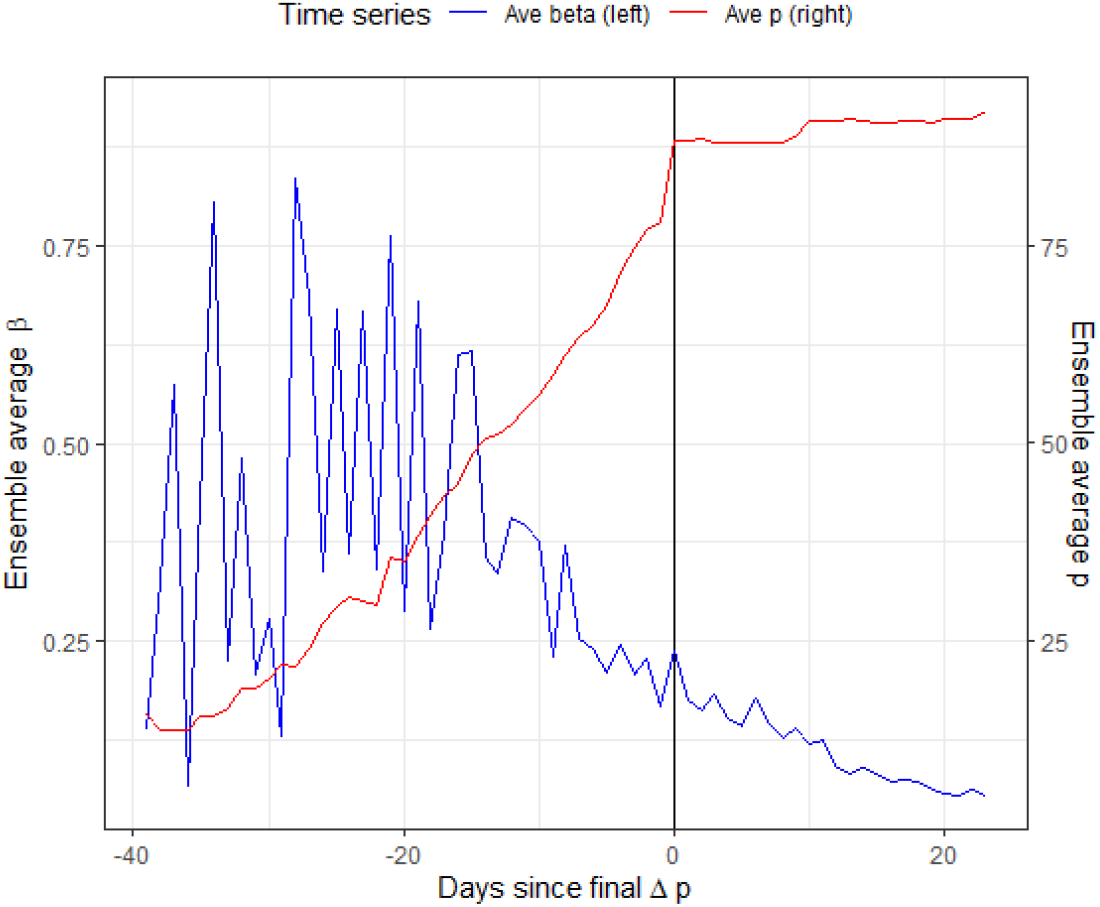
Comparison of ensemble average NPI stringency index, *p* against the ensemble average of *β* as a function of days from the last major change in control value.

**Figure 6:**
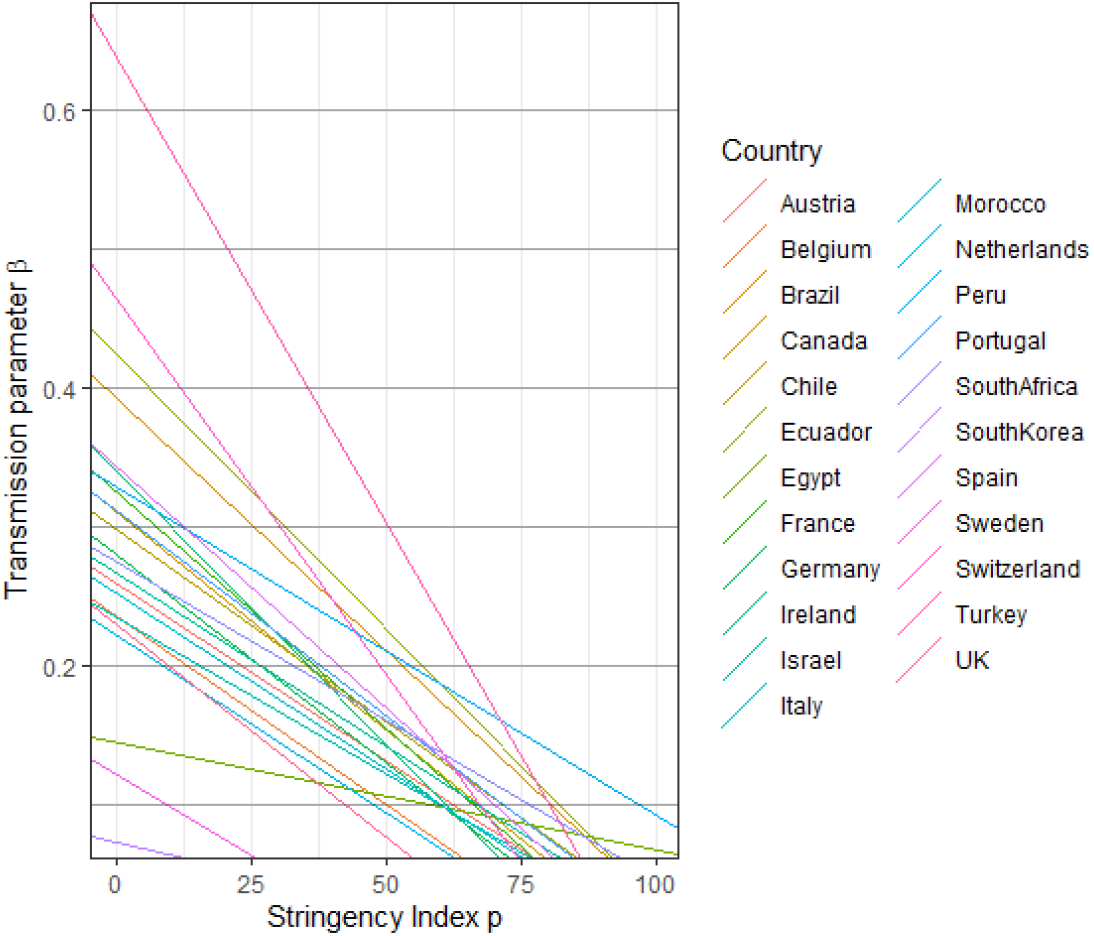
Control surfaces of the model on observed transmission rate and stringency index for ensemble of countries within the study.

**Figure 7:**
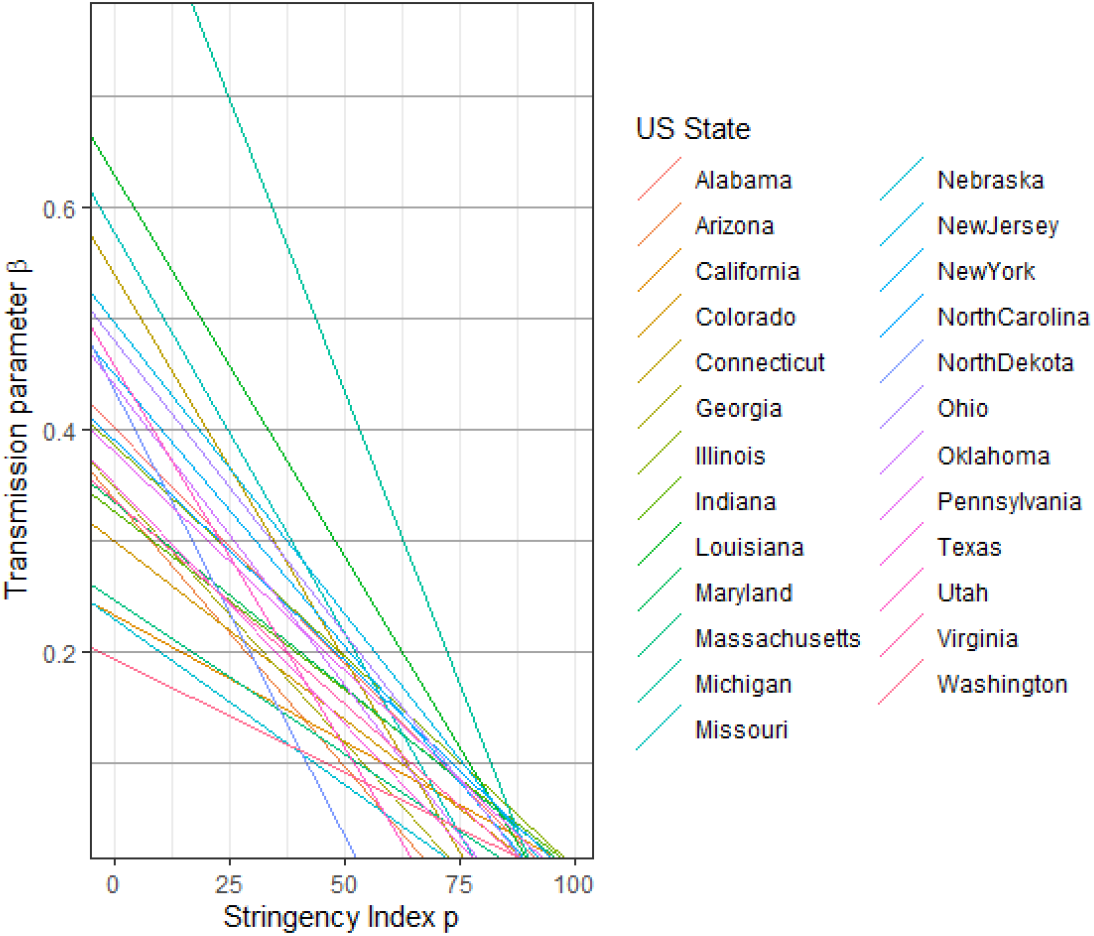
Control surfaces of the model on observed transmission rate and stringency index for ensemble of states within the United States.

## 5. Discussion

One of the most important results from Tables 2 and 3 is that, for the bulk of countries and US States, the intervention leverage *α_s_* ≈ 0.01. It is remarkable that this occurs whilst other parameters are measured to have a great variety, especially *β*_0_. This striking result also speaks to the universal character of the stringency index used here to quantify NPIs.

The next most important result is the universal time domain response of the transmission rate, *β* to a change in stringency of control as depicted in Figure 5. The ensemble averages of *β* and *p* produce a very clear exponential decay to enhanced control which is well modeled by the kernel function proposed in eq. (1). For visual confirmation, a representative sample of countries are depicted in Figure 8.

**Figure 8:**
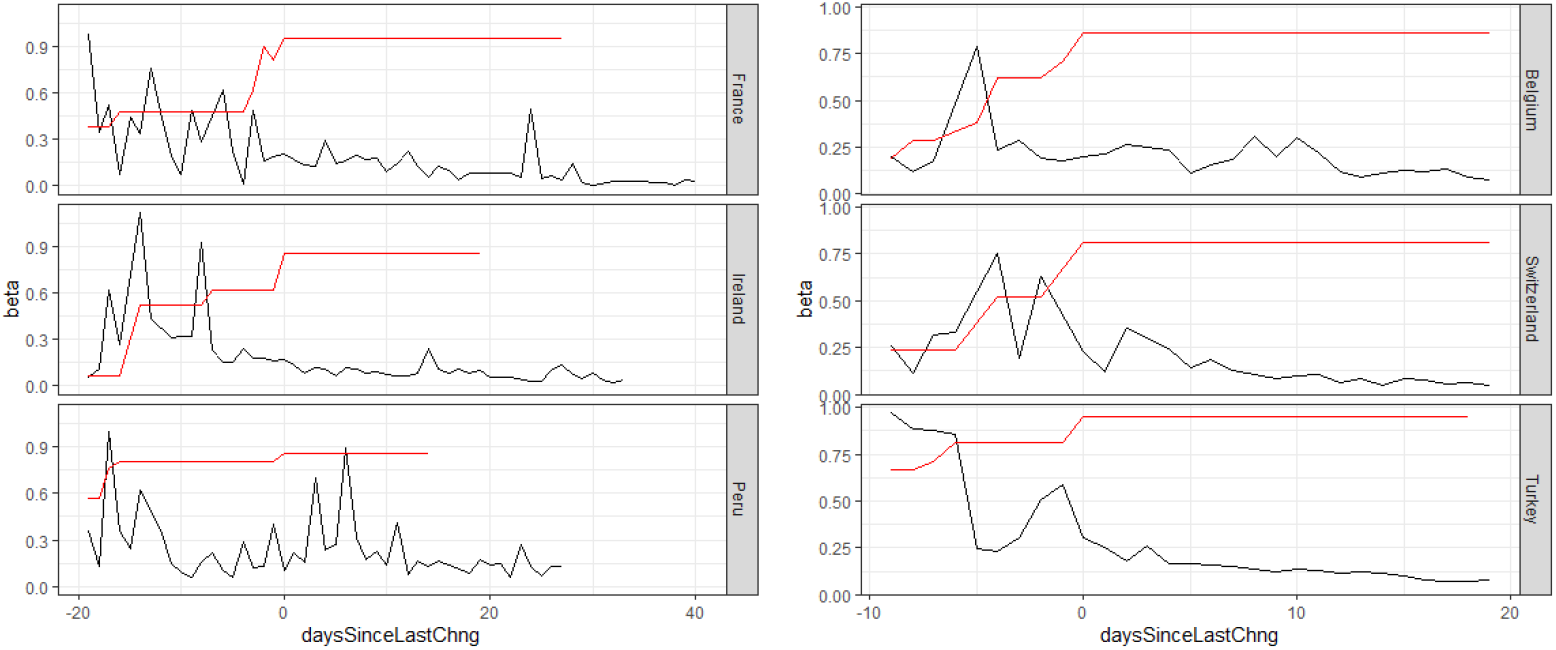
Representative sample of transmission rates in response to NPIs for countries. The exponential kernel in the response (black) to change in NPI (red) is clearly evident in all of these.

### 5.1. Predictive capabilities of the model

The recent few weeks of extreme containment measures across the world have yielded valuable data to allow for country-specific predictions. With respect to transmission rate, our model can answer *how much?* change we can expect to see and *by when?* we expect to see it as function of p; as seen in eq. (5). This was derived using eqs. (1) and (2). Hence,

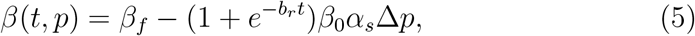

where *t* = 0 corresponds to the end of the lockdown period and the enacting of new interventions, such that *p* < *p_max_*. The next few weeks of post-lockdown actions by a number of countries and states may refute this model and should be watched with interest.

### 5.2. Predictions for Italy

For illustration purposes, we choose the configuration of parameters obtained with Italian data, where *ϕ* = 0.1 is used (see Section 3.2). The scenario considered here assumes that variations of the index occur after the last available data with Δ*p* = −10, −30.

Figure 9 depicts the time dependence of the number of symptomatic population, the active cases and the cumulative number of cases and fatalities. One can appreciate that going from Δ*p* = −10 to Δ*p* = −30 has serious consequences in terms of the time it takes to achieve the peak in active case as well as the amplitude of the peak.

**Figure 9:**
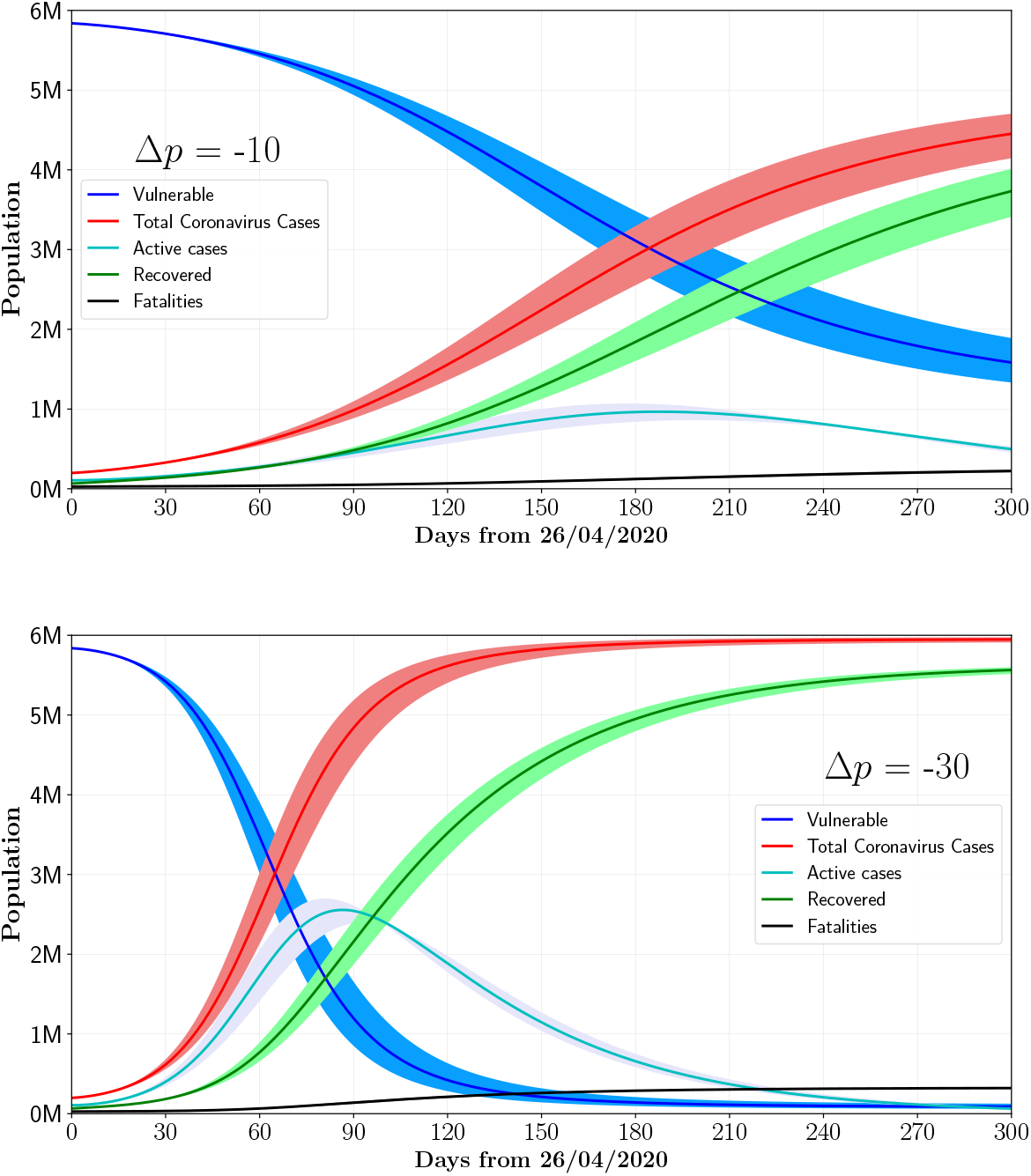
Post-lockdown scenarios for Italy for *p* = 85 (upper plot), *p* = 65 (lower plot). Results are given using *ϕ* = 0.1 of the susceptible population population, or vulnerable population, active cases and the cumulative distribution of total cases and fatalities. The bands correspond to the uncertainties in the model (see text).

This is further illustrated in Figure 10, where the number of active cases and daily fatalities are shown as a function of time for different Δ*p* ranging from −10 to −50. The time required for the peak to occur would go from over six months to about four and the amplitude of the peak would double for a relaxation of Δ*p* = −20 instead of Δ*p* = −10. The uncertainty envelope in these predictions is driven primarily by the ensemble variation of *α_s_*, which is of order of 10%. This is considered to be a more realistic estimate of the potential deviation from the linear behavior assumed in eq. (2). The other source of uncertainty is the daily recovery, *γ*, which is much smaller. Graphs in Figure 9 display the bands that incorporate these uncertainties.

**Figure 10:**
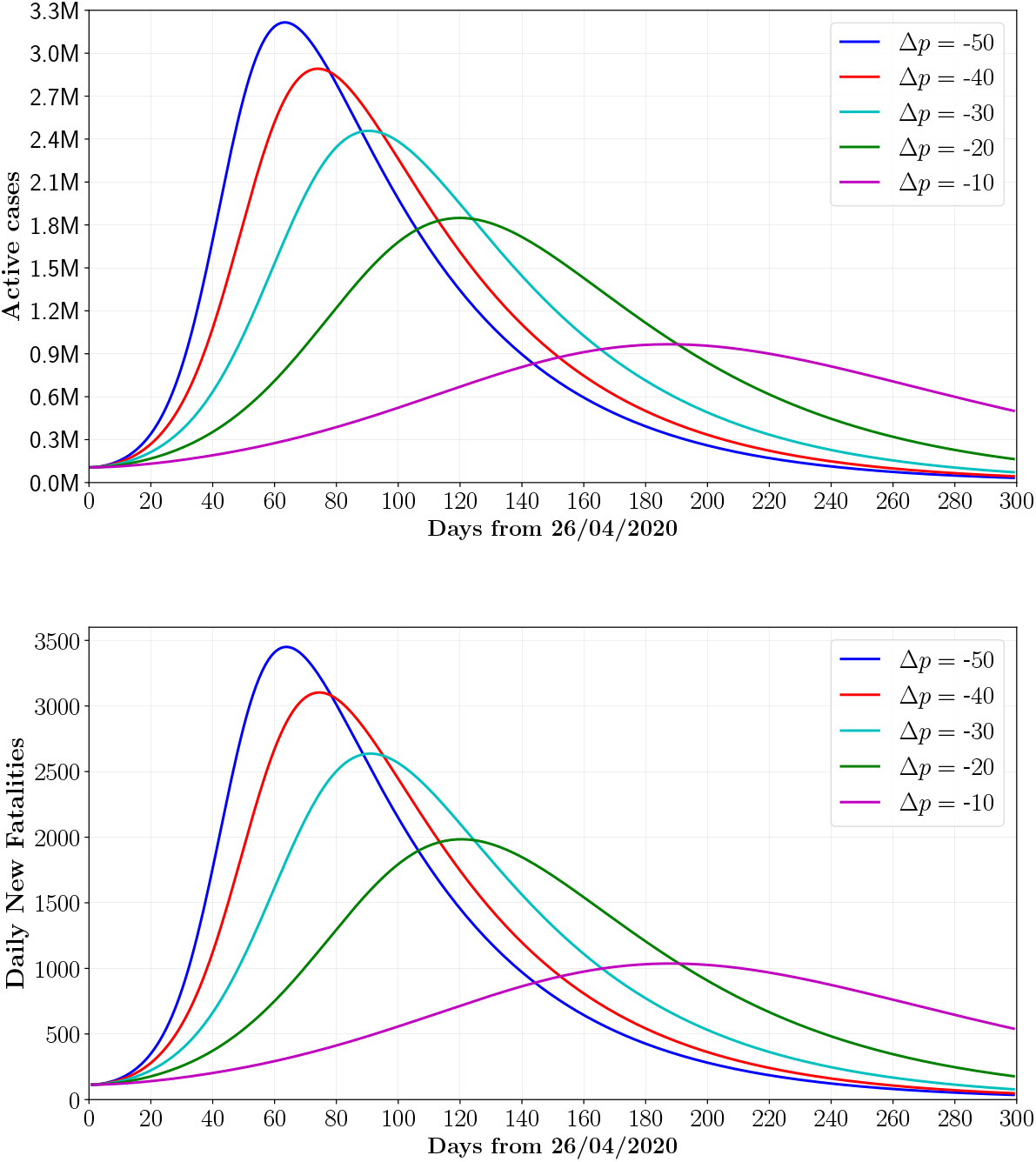
Number of active cases and new fatalities as a function for time for Italy by assuming different values of Δ*p* ranging from −10 to −50. Results are computed with *ϕ* = 0.1.

Figures 9 and 10 predict that releasing containment measures too swiftly would lead to the resurgence of very large peaks of symptomatic infections within a month or two, leading to even worse outcomes compared to those observed so far.

### 5.3. Main synthesis: acceptable NPI control levels

Our entire synthesis regarding the value of NPIs as a measure of control of the current pandemic is captured by Figure 11. Once you define the concept of *criticality*, this synthesis follows by deduction.

**Figure 11:**
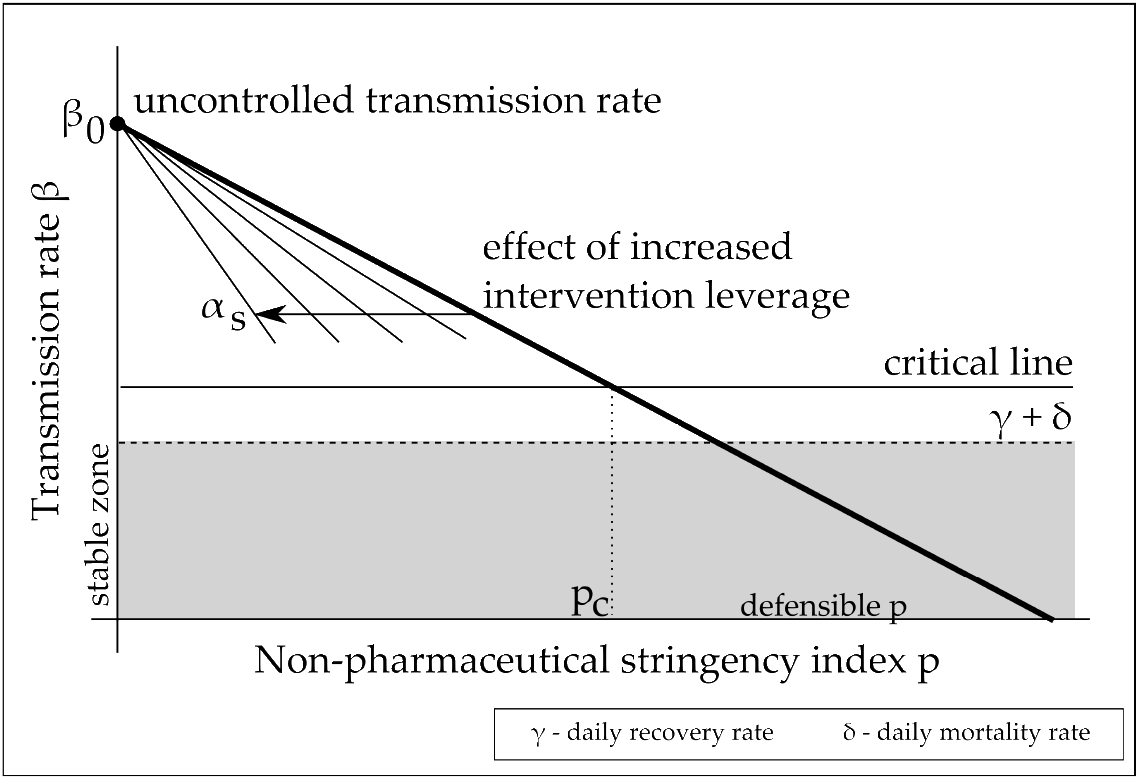
The constraints on the NPI control surface as it relates to the transmission rate. The intervention leverage, *α_s_* affects how easy it is for a country to reduce transmission rate for a given stringency index level. There is a linearly stable zone, meaning that the observed daily transmission rate is less than the combined daily recovery *γ* and daily death rate *δ* i.e. *R_t_* < 1. By definition, the critical level of NPI *p_c_* may not be reduced further or there will be a guarantee of overwhelming the healthcare system. Note that the critical line moves upward as the size of the susceptible population decreases.

#### Definition

Criticality is the allowable daily transmission rate, given the current daily recovery rate and daily death rate such that some constraint is almost, but not quite, violated.

For illustration, consider the constraint of keeping peak active cases below a threshold to prevent the country’s health care system from being overwhelmed. There exists a critical line in the control surface of Figure 11, for a given *γ* + *δ*, that achieves this uniquely for that locale.

It is a very general principle, that the critical line is raised as the susceptible population is reduced in number. The full non-linear model in eq. (C.1) demonstrates this, as fewer and fewer people are susceptible the lower the need for NPIs to remain sub-critical.

If the desire is to relax NPIs below the current critical NPI level, *p_C_*; the only other degree of freedom left is the intervention leverage *α_s_* which will need to be closely monitored. Outlier locales in Tables 2 and 3 have demonstrated that this is possible.

### 5.4. Monitoring intervention leverage is important

It is paramount to closely monitor the evolution of intervention leverage *α_s_* as NPIs are released. If the clinical situation does not change, *γ* and *δ* will remain small. The allowable transmission rates will also have to remain small as well and therefore strict measures of NPIs will need to remain in place.

It is essential to increase *α_s_* if the desire is to relax NPIs beyond the current critical NPI level, *p_c_* in Figure 11. By increasing *α_s_*, the critical NPI level is ‘left shifted’ and allows for an even greater relaxation of NPIs than would otherwise be possible. For the system to remain sub-critical, the rate with which *α_s_* increases should outpace that of the decrease of the stringency index if a total relaxation of NPIs is desired. Monitoring of *α_s_* becomes essential to controlling the post-lockdown phase when controlled in this regime. For further details, see Appendix E.

### 5.5. Easier compliance with softer NPIs

One is tempted to speculate about the possibility of non-linear behavior in Eq. 2, where data would favor *α_s_* to increase as *p* decreases i.e. it would be easier for citizens to comply with less stringent NPIs. This would be good news in terms of the effort required to manage the pandemic in that the effectiveness of containment measures could increase as *p* decreases. This argument is hindered by the fact that the lapse of time between changes in the observed non-pharmaceutical interventions was not significant and measuring the effect of each individual intervention was therefore difficult. As a result of swift action by Governments, we observe the effect of an ensemble of more or less stringent measures, as opposed to their sequential application. To this end, we lack the evidence that would support the above mentioned non-linear behavior.

### 5.6. Insights from Outliers

It is appropriate to comment on some of the outliers identified in the estimation of intervention leverage *α_s_*. Table 4 summarises this discussion; density of people plays a key role in intervention leverage and a correlation study with World Bank data substantiates this claim further (See Appendix D for further details). It need not be ignored that the prolonged spread of the virus during the early stages of the pandemic in Italy and Spain were driven by lack homogeneity in the adherence to advisories. This prompted governments to introduce severe restrictions to movement, where law enforcement agencies became heavily involved in ensuring compliance. The importance of public awareness and compliance with regards to NPIs to slow the spread is hence embodied by the intervention leverage *α_s_* and is an important observable during the post-lockdown stage.

**Table 4:** Locales with relatively large *α_s_* values and low *P_max_* relative to other states and counties in the study set.

| Locale | $p_{max}$ | $\alpha_s$ | No. cities pop. greater than 1M |
| --- | --- | --- | --- |
| Arizona | 62 | 0.014 | 1 (Phoenix, 1.6M) |
| North Dakota | 48 | 0.018 | 0 |
| Utah | 62 | 0.015 | 0 |
| Sweden | 52 | 0.019 | 1 (Stockholm, 1.6M) |
| Switzerland | 81 | 0.012 | 0 |

### 5.7. Recommendations

A world wide survey paper is intended to follow our results where the key parameters of our latent SIRD model are correlated with macro-economic, public health and geo-physical measurements to determine the environmental effects on the transmission, recovery and mortality parameters of COVID19. While lockdown measures have been successful in curbing the spread, our study indicates that removing them too swiftly will result in the resurgence of the spread within one to two months. Reducing the stringency index by 10 will delay reaching the apex by about 6 months, where reducing it by 20 will delay by only four. The peak cases almost double by moving from Δ*p* = −10 to Δ*p* = −20. This indicates that post-lockdown measures should be staged and the reduction of the stringency index should be slow.

## Data Availability

All data incorporated is publicly available.

https://coronavirus.jhu.edu/map.html

## Contributors

JC - Data collection, data analysis FC - Data analysis, review of writing SD - Data analysis, simulation, figures BD - Data collection, review of writing LD - Data collection, review of writing KH - Data collection, Data analysis BL - Data collection, data analysis, figures, tables CM - Literature search, writing review BM - Study design, writing, data interpretation, data analysis, modeling KM - Data collection, data analysis, figures JN - Literature search, figures, writing, data analysis, data interpretation, modeling XR - Data analysis FS - Data collection, data analysis, figures, writing

## Declaration of interests

All authors declare no competing interests.

## Acknowledgements

Authors are indebted to the South African Department of Science and Innovation and the National Research Foundation for different forms of support. This includes, but it is not limited to, support through the SA-CERN Program and the National E-science Postgraduate Teaching and Training Platform. Authors are also grateful for grant support from the IEEE. The funders of this project had no role in study design, data collection, data analysis, data interpretation, or writing of the report. BM, JN, BL, FS, SD, JC, BD, LD, KH, KM, XR had access to all of the data. The corresponding author had final responsibility for the decision to submit for publication.

## Appendix A. Stringency Index for United States of America

For each policy response measure S1-S7, OxCGRT use the ordinal value (and add one if the policy is general rather than targeted). This creates a score between 0 and 2 and for S5, and 0 and 3 for the other six responses^29^.

The OxCGRT stringency index is given by:

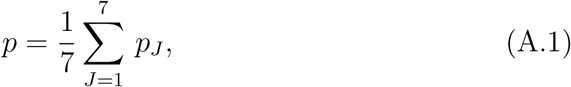

where *p_J_* is defined by:

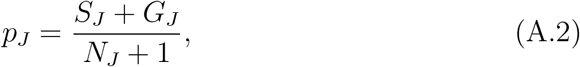

with *G_J_* = 1 if the effect is general (and 0 otherwise), and *N_J_* is the cardinality of the intervention measure^29,30^. In the case where there is no requirement of general vs. targeted (S7), the +1 in the denominator and the *G_J_* in the numerator are omitted from the equation to form:

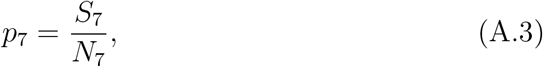

The OxCGRT database contains data for 133 countries however it does not contain specific data for US states. It is important to be able to compare the US states non-pharmaceutical interventions (NPIs) with those of other countries around the World in a unified framework.

To this end, we coded the known levels of intervention in America to match as nearly as possible, the OxCGRT system. We used the Institute for Health Metrics and Evaluation (IHME) dashboard to obtain six dates at which specific states imposed different NPIs^31^.

**Table A.5:** Table Showing US Interventions Acquired from the IHME.

| Label | NPI |
| --- | --- |
| $U1$: | Stay at Home Order |
| $U2$: | Educational Facilities Closed |
| $U3$: | Non-essential Services Closed |
| $U4$: | Travel Severely Limited |
| $U5$: | Initial Workplace Closure |
| $U6$: | Banned Mass Gatherings |

In order to compare the US intervention data it was necessary to make a stringency index for the US states that mimics that of the index that was made for the World data by OxCGRT.

The following decisions were made during the process of mapping the reported US NPIs to the OxCGRT index:

- The US *Mass Gatherings Banned U*5 can be mapped directly to the Oxford *Cancel Public Events S*3.
- The US *Initial Business Closure U*6 can be mapped to the Oxford *Work Place Closure S*2
- The US *Travel Severely Limited U_4_* can be mapped to the Oxford *Domestic Travel Bans* S6 and *International Travel Bans* S7 combined.

**Table A.6:** Table Showing Unmapped NPIs.

| Label | Interventions Not Directly Interchangeable |
| --- | --- |
| US NPIs: |  |
| $U1$: | Stay at Home Order |
| $U3$: | Non-essential Services Closed |
| OxCGRT NPIs: |  |
| $S4$: | Close Public Transport |
| $S5$: | Public Information Campaign |

Although some of the above US interventions were not directly comparable to the OxCGRT indicators, their individual impact on the stringency index is still valid and should be included in the index. By including *U*1 and *U*3 with the appropriate weight into the same calculation OxCGRT used for their index, an equivalent US index is created. The following equation was developed:

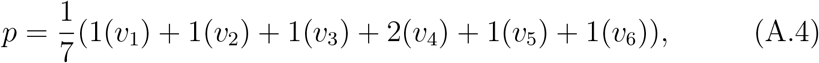

where *v_i_* is a number out of 100 indicating the extent each of the interventions are imposed.

Due to lack of data on the *Travel Severely Limited* intervention in the IHME database. It was necessary to source US travel restrictions information from other US news sources. There have been a number of travel interventions that have been implemented however there have not been widespread travel bans between states. ^32^ The first significant travel restriction was a US-Europe travel ban to 26 European countries, which was announced on 11 March 20 20.^33^ On 19 March 2020 the US issued a level 4 “Do not travel” advisory which is the highest travel restriction in the US. US citizens were informed that they can travel back to the US if they were out of the country when the ban was announced but if they do not do so timely they might find themselves having to stay abroad for an extended period of time. Foreign nationals who have been to the 26 EU countries or the UK, China, Iran or Ireland are not allowed entrance to the US.^34^ In terms of the interstate travel restrictions. There have been no full travel bans but in some states you are required to quarantine for 14 days after arrival.^32^ Therefore, it was necessary to introduce a leveled implementation of the U4 *Travel Severely Restricted* measure. Using the same logic used by the Oxford COVID-19 Government Response Tracker (OxCGRT) the following equation for *v_i_* was introduced:

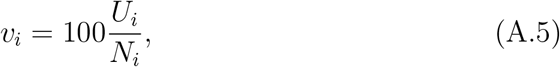

where *U_i_* is ordinal and can vary from 0 to the cardinality of the intervention measure, *N_i_*. This is for the purpose of incorporating levels of implementation of specific interventions into the stringency calculation. Based on the data the only intervention that requires levels of implementation is the *U*_4_ intervention.

The following ordinal levels were employed for *U*_4_:

0. No travel restrictions (US before the 11 March 2020)
1. US preliminary travel ban to 26 EU Countries (Commenced 11 March 2020)
2. Level 4 “Do not travel” advisory issued (19 March 2020)
3. Interstate Travel bans (No interstate travel bans are currently imposed)

The specification for comparable *p* among countries within the OxCGRT database and the United States of America is completed with the above definitions.

## Appendix B. Model Details

An explanation of the model is warranted. It has a causal structure and may be clinically interpreted as well, both of which are desirable properties for a model which needs to be controlled. The causal structure gives insight into *what* to control and the clinical interpretation gives insights into *how*.

### Susceptible individuals, S

These are the unexposed and susceptible individuals within the population and include healthcare workers as well general members of the public.

### Observed infections, I

These infections represent patients who have tested positive for COVID-19 and are actively reported on^10^. Any contacts of these patients who subsequently test positive, any nosocomial infections due to these patients (for example, healthcare workers who contract the disease) or any other individuals who knowingly interact with COVID-19 positive patients are modeled by *β*, the transmission rate of COVID-19 amongst observed infections. Well prepared countries with strict healthcare protocols for known positive patients, for example quarantining, effective use of personal protective equipment for healthcare providers, and physically separate care pathways for positive patients all essentially work to ensure that *β* is kept as small as possible.

Asymtpomatic also transmissable Some of these observed infections, *I* are due to some mild or asymptomatic cases which become severe enough to warrant testing or cause patients to seek medical attention. These cases are modeled by *ϕI_L_*, where *ϕ* is dependent on the probability that a latent infection *I_L_* becomes a known positive case, *I*. Latent infections are addressed next.

### Latent infections, I_L_

There is evidence of a non-trivial fraction of cases going undetected as a result of presenting with mild symptoms or being asymptomatic^35,36^. This is the reason for including the latent variable dynamics within our modification of the standard SIRD model.

Table 1 in the main text has good estimates of asymptomatic cases; together with the patients who have subclinical manifestations of Covid-19, these cases are all included in the latent infection group, *I_L_*.

It is the susceptible group’s interaction with these asymptomatic and mild cases which produce new latent infections and this is modeled through *β_L_*, the non-negligible latent transmission rate.

### Latent recoveries, R_L_

A majority of these asymptomatic and mild symptom patients resolve the virus using their natural immunity without ever being tested. Early reports indicate that this may be a substantial number of latent infections^16,18^. These cases eventually form part of the latent recovered group, *R_L_*. The rate of recovery of the latent infected group is captured by *γ_L_*.

### Latent infections dying, *δ_L_I_L_*

These counts are considered weakly observable and rare. The weak observability is present with the revised, erratic death counts by some officials when home visits uncover additional cases^10^. It is unclear that these may be directly attributable to the virus in that confirmation would be required via post-mortem COVID-19 testing?. Given the existing testing burden posed on most countries, this sort of testing is rare and, therefore, the uncertainty in this parameter will remain high^18^.

### Known recoveries, R

These are patients who are known to test positive for COVID-19 and are known to have recovered fully from the virus. The recovery rate, *γ*, models how quickly known infections are resolved and discharged out of the healthcare system. This rate is physically dependent on treatment regime and the patient’s own physical condition.

### Deceased patients, D

The number of deceased individuals is denoted by *D*; and it is mostly affected by the known individuals who have tested positive and are currently being treated in the prevailing healthcare system. The implications for the model are that *δ* >> *δ_L_* i.e. under normal treatment and monitoring situations, the implied probability of fatality for a known infection is much larger than the implied probability of fatality for a latent infection.

Under the above conditions, the model explicitly caters explicitly for the situation that the latent infections are asymptomatic or mild.

It is trivial to show that *S* + *I* + *I_L_* + *R_L_* + *R* + *D* = *N* at every instant in time. Furthermore, the model in Figure 1 implies that:

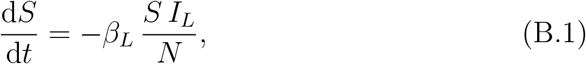

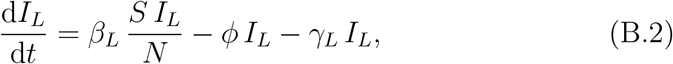

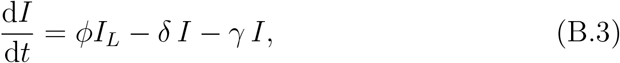

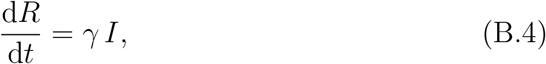

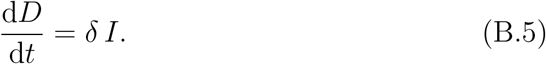

### Appendix B.1. Linearisation

Each of the equations in (B.1) - (B.5) were linearised using the approximation that *S* = *N* – *ϵ*, which gives equivalent results to the Jacobian method^27^. This operating point corresponds to the case where the epidemic is still in the early/controllable phase and the number of infected individuals is small compared with the size of the total population *N*.

The derivatives with respect to time were approximated using first order backward difference approximations at a daily level. Classical frequentist error propagation was applied to this linear approximation using the Gaussian process assumption.

Theoretically, these approximations are valid provided that:

1. the number of infected are a small fraction of the susceptible population.
2. the dynamics of the disease process are slow compared with a single day. This is justified by the work from Weiss and Murdoch^36^.

The final form, used for analysis of the time variation of the parameters as various forms of control are applied is:

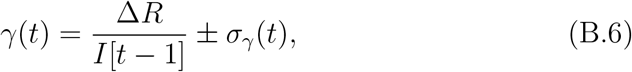

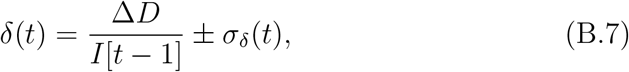

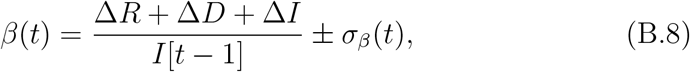

where *σ_x_*(*t*) is the noise estimate at day *t*, Δ*x*: = *x*[*t*] – *x*[*t* – 1] and *x* is either *R*, *D* or *I*.

### Appendix B.2. Error propagation

Using the coefficient of variation and propagating the error in the linear approximation, assuming that the errors in the daily counts are within 10%^37^, the probable noise levels in the daily time variations are calculated by:

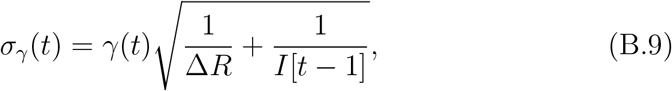

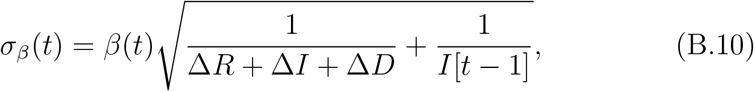

and

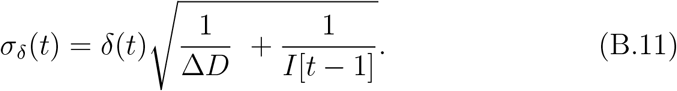

These results depend on the count data being Poisson processes and the fact that the coefficient of variation of a Poisson process is λ^−1/2^. Recall that the general coefficient of variation of a division of two random variables is the quadrature sum of the numerator and denominator coefficients of variation; the results follow ^37^.

### Appendix B.3. Details of Linearisation

If d*t* is taken as one day, *t* is the day index and Δ*x* = *x*[*t*] – *x*[*t* – 1], then with the modeling assumptions the differential equations simplify to:

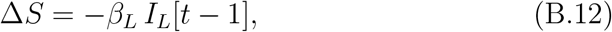

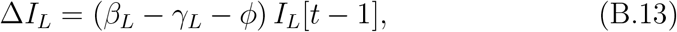

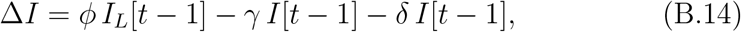

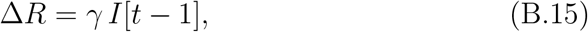

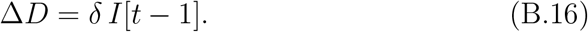

Use of (B.15) and (B.16) yield the daily estimates of the observable recovery rate, *γ* and fatality rate *δ*. Combining (B.15), (B.16) and (B.14) give the daily estimate of the transmission rate, as observed through the detection efficiency.

## Appendix C. Control Dynamics

Efforts to control the pandemic do not happen everywhere, all at once and this is the reason that the control efforts can be said to be dynamic. Indeed, the form of the kernel functions in equations (1) - (4) state this implicitly. Each of the observed parameters will be looked at it in this section and their dynamics described. These forms have non-trivial implications for control of the pandemic and will be expounded upon in a follow up paper.

### Transmission rate

The beta kernel in (1) and the steady state behaviour modeled by (2) imply that the transmission dynamics, under stringency of control p, behave as a first order control system^27^:

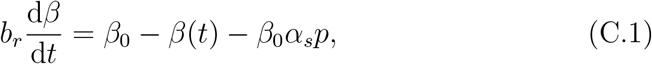

where *b_r_* is the typical adjustment time for a control measure to take full effect, *α_s_* is the societal sensitivity (estimated for each country in our work) to control measures *p* ϵ [0,100], and *β*_0_ is the uncontrolled transmission rate within a society.

As a sense check; when *p* = 0 then the equilibrium condition of (C.1) is found by solving (C.1) with *β*(*t*) = *β*_f_ = *const*. The non-trivial solution is *β_f_* = *β*_0_ and shows that, without control, the transmission rate becomes *β*_0_. This is defined as the uncontrolled transmission rate and is as it should be.

If *p* ≠ 0, then the equilibrium condition is *β_f_* = *β*_0_ – *β*_0_*α_s_p* which is exactly equation (2). The general analytic solution to (C.1) is precisely the kernel function in (1).

It is this form which allows for the use of classic and modern control methods to shape *β*(*t*) to a form that is acceptable for the desired goals of the pandemic control system eg. minimise total deaths, minimise the peak load on the health care system, maximise the economic activity etc. These aspects will be dealt with in detail in a follow up paper.

## Appendix D. Correlation of transmission rate with macroscopic indexes

The phases of the spread and its parameters bear strong similarities in a wide range of countries considered here. However, non-trivial differences in terms of parameters can be observed when scrutinising country by country variations. It is relevant to correlate variances with respect to macroscopic indexes. For this purpose the number positive cases is analysed as a function of time using the parametric expression:

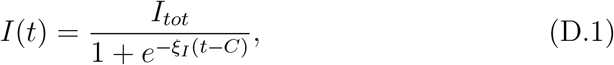

where *I_tot_* denotes the total expected number of positive cases for *t* → ∞, *ξI* is the slope of the exponential growth that characterises the first phase of the spread, and *C* can be interpreted as a measure of the time needed to deviate from the initial exponential growth leading to containment. Time is expressed in number of days. A total number of 67 countries are selected, where containment measures have proven effective in curbing the spread. These include countries in all continents and with a wide span in terms of socio-economic development, inequality and population density.

Macroscopic indicators are organised according to relevant themes: socioeconomic vulnerabilities, demographics, social expenditures and aggregate economic indicators. A total of 34 indicators from the World Bank data base are selected and are correlated with the parameter £/ from each country.

The Gini index quantifies the extent to which the distribution of income among individuals or households deviates from perfect equality. The selected sample of countries displays a minimum and a maximum Gini index of 24 and 50, respectively. It is found that the parameter *ξI* is almost insensitive to the Gini index. This is illustrated in Figure D.12 where the red line corresponds to a first order polynomial that is consistent with zero slope. In order to exclude statistical fluctuations in the sample a similar study is performed using the percentage share of income held by the lowest 10% and 20% of the income bracket. No significant correlation is found for either of the indexes. This indicates that social inequality is not strongly correlated with the rate of spread of the virus.

**Figure D.12:**
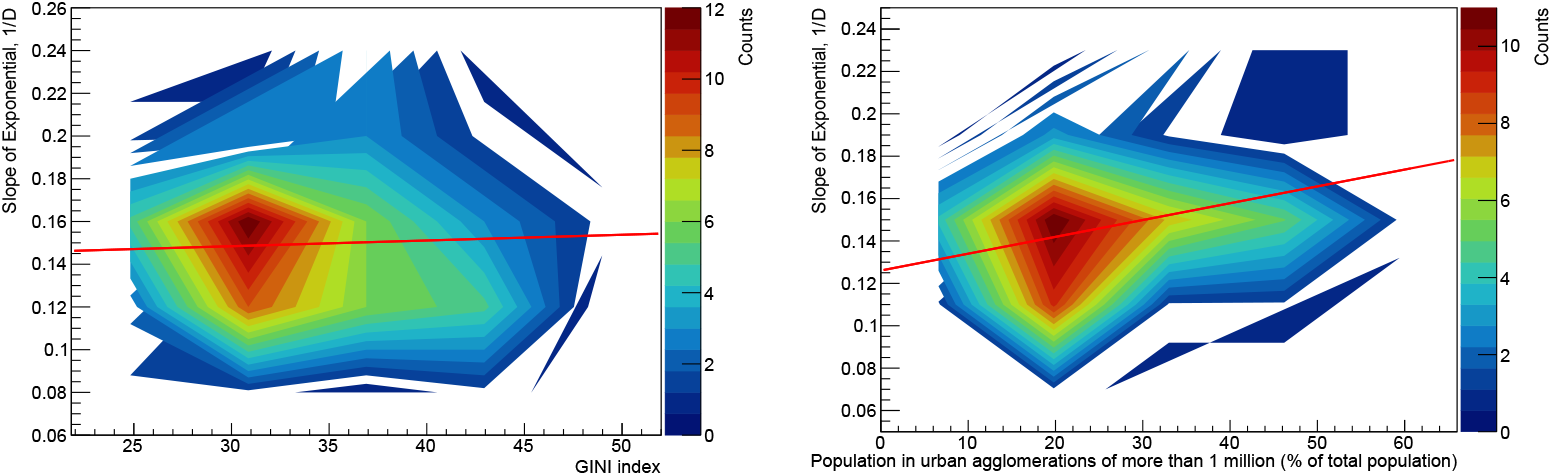
Correlation of *ξI* (see text) with the Gini index (left) and the population in urban agglomerations of more than one million to the country’s population living in metropolitan areas (right). The red lines correspond to first order polynomials that illustrate the degree of correlation.

This observation is further strengthened by evaluating the correlation with the proportion of the urban population living in slum households. According to the World Bank, a slum household is defined as a group of individuals living under the same roof lacking one or more of the following conditions: access to improved water, access to improved sanitation, sufficient living area, and durability of housing. Out of the 67 countries under scrutiny, 18 report a significant fraction of urban population living in slums. In this sample of countries the fraction ranges from 8% to 53%. No significant correlation is found between this index and *ξI*. In addition, the average value of *ξI* for these 18 countries is compatible with that of the rest of the ensemble studied.

As per the physical picture underlying the model used to describe the spread, it is expected that population density should play a significant role. No significant correlation is found with the average population density. This can be explained by the fact that the average population density is not necessarily a good metric for population density in urban areas, where the spread is most likely to occur. It should be noted that the correlation with the fraction of the population in urban areas is not statistically significant. In order to scrutinise the relevance of localised population density, the index made of the population in urban agglomerations of more than one million to the country’s population living in metropolitan areas in percentiles is used, as illustrated in Figure D.12. It is found that the correlation can be parametrised with a first order polynomial with a slope of (1 ± 0.3)·10^-3^ per day. The significance of the correlation greater than a 3 σ Confidence Level. This the most significant correlation out of all the indexes considered here.

## Appendix E. Inequality for relaxing beyond the critical NPI level

It will be argued that one of the strategies for the post-lockdown period is to increase intervention leverage *α_s_* as *p* decreases. This will sustain quasilinear behavior in the number of active cases and it is depicted in Figures 9 and 10 for Δ*p =* −10.

In practice, all relevant parameters that enter the temporal evolution described above need to be tuned such that the expected peak of the number of cases and ICU usage falls below the thresholds characteristic to each country. One can denote a vector of critical parameters for which a country’s healthcare system is not overwhelmed:

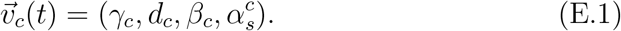

This is what is meant by *criticality*; the value of parameters such that the healthcare system is only marginally not overwhelmed. One can assume that *γ_c_* and *δ_c_* display a weak time dependence in that they primarily depend on medical advances, rather than on NPIs. In this setup the condition for the system to remain sub-critical can be expressed as follows:

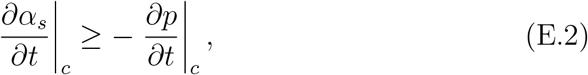

where the temporal partial derivatives are evaluated at the point of criticality defined by 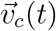. The inequality (E.2), while seemingly straightforward from a mathematical standpoint, has serious consequences for policy makers. While lockdown measures have been successful in bringing the reproductive factor down to one and below, it is evident that these are having devastating effects on the economic landscape. In African countries, lockdown measures necessary to control the epidemic are leading to widespread malnutrition in vast sections of the population.

On the other hand, the illustrative example shown in Section 5.1, indicates that for fixed intervention leverage *α_s_*, reducing *p* significantly can lead to the advent of an epidemic of unprecedented proportions. Under these conditions, the inequality (E.2) speaks to the need to ensure that the rate with which intervention leverage *α_s_* grows should outpace that of easing non-pharmaceutical interventions.

## Notes

### Competing Interest Statement

The authors have declared no competing interest.

### Funding Statement

South African Department of Science and Innovation
South African National Research Foundation
SA-CERN Program
National E-science Postgraduate Teaching and Training Platform
IEEE

